# Antibody dynamics to SARS-CoV-2 in asymptomatic COVID-19 infections

**DOI:** 10.1101/2020.07.09.20149633

**Authors:** Qing Lei, Yang Li, Hongyan Hou, Feng Wang, Zhuqing Ouyang, Yandi Zhang, Danyun Lai, Banga Ndzouboukou Jo-Lewis, Zhaowei Xu, Bo Zhang, Hong Chen, Junbiao Xue, Xiaosong Lin, Yunxiao Zheng, Zongjie Yao, Xuening Wang, Caizheng Yu, Hewei Jiang, Hainan Zhang, Huan Qi, Shujuan Guo, Shenghai Huang, Ziyong Sun, Shengce Tao, Xionglin Fan

## Abstract

**Importance:** Asymptomatic COVID-19 infections have a long duration of viral shedding and contribute substantially to disease transmission. However, the missing asymptomatic cases have been significantly overlooked because of imperfect sensitivity of nucleic acid testing. We aimed to investigate the humoral immunity in asymptomatics, which will help us develop serological tests and improve early identification, understand the humoral immunity to COVID-19, and provide more rational control strategies for the pandemic.

**Objective:** To better control the pandemic of COVID-19, dynamics of IgM and IgG responses to 23 proteins of SARS-CoV-2 and neutralizing antibody in asymptomatic COVID-19 infections after exposure time were investigated.

**Design, setting, and participants:** 63 asymptomatic individuals were screened by RT-qPCR and ELISA for IgM and IgG from 11,776 personnel returning to work, and close contacts with the confirmed cases in different communities of Wuhan by investigation of clusters and tracing infectious sources. 63 healthy contacts with both negative results for NAT and antibodies were selected as negative controls. 51 mild patients without any preexisting conditions were also screened as controls from 1056 patients during hospitalization in Tongji Hospital. A total of 177 participants were enrolled in this study and serial serum samples (n=213) were collected. The research was conducted between 17 February 2020 and 28 April 2020. Serum IgM and IgG profiles of 177 participants were further probed using a SARS-CoV-2 proteome microarray. Neutralizing antibody responses in different population were detected by a pseudotyped virus neutralization assay system. The dynamics of IgM and IgG antibodies and neutralizing antibodies were analyzed with exposure time or symptoms onset.

**Results:** Asymptomatics were classified into four subgroups based on NAT and serological tests. In particular, only 19% had positive NAT results while approximately 81% detected positive IgM/IgG responses. Comparative SARS-CoV-2 proteome microarray further demonstrated that there was a significantly difference of antibody dynamics responding to S1 or N proteins among three populations, although IgM and IgG profiles could not be used to differentiate them. S1 specific IgM responses were elicited in asymptomatic individuals as early to the seventh day after exposure and peaked on days from 17d to 25d, which might be used as an early diagnostic biomarker and give an additional 36.5% seropositivity. Mild patients produced stronger both S1 specific IgM and neutralizing antibody responses than asymptomatic individuals. Most importantly, S1 specific IgM/IgG responses and the titers of neutralizing antibody in asymptomatic individuals gradually vanished in two months.

**Conclusions and relevance:** Our findings might have important implications for the definition of asymptomatic COVID-19 infections, diagnosis, serological survey, public health and immunization strategies.

## Introduction

SARS-CoV-2 is an emerging coronavirus, which was first recognized as the causal agent of COVID-19 in December 2019,^1^ and has rapidly spread around the world. On 11 March 2020, the WHO has declared COVID-19 a global pandemic.^2^ As of 28 June 2020, there have been 9,843,073 confirmed cases and 495,760 deaths, reported in 215 countries and territories worldwide.^3^ Unlike the epidemic of SARS-CoV and MERS-CoV, the sharp rise in the number of global COVID-19 cases in a short epidemic episode brings the fear of having viral transmission from asymptomatic individuals. On Jan 28, 2020, the National Health Commission of China updated the COVID-19 Prevention and Control Plan (3th edition) and first emphasized the identification and quarantine of asymptomatic infections.^4^ Asymptomatic COVID-19 infection has been defined as a person infected with SARS-CoV-2 who has no clinical symptoms (such as fever, cough, or sore throat, etc.) and no radiological changes of the lung, yet nucleic acid testing (NAT) positive for SARS-CoV-2.^4^ As of February 11, 2020, there were 72,314 COVID-19 cases reported in China, and 889 cases (1.2%) belonged to asymptomatic infections.^5^ As of April 14, 2020, a total of 6,764 asymptomatic infections reported in China, which accounts for about 5.9% of all cases. Among these, 1,297 asymptomatic individuals, in fact presymptomatics and subsequently developed to the confirmed cases with different severities, while the others remain asymptomatic.^6^ Moreover, several studies reported that asymptomatic COVID-19 individuals play an important role in the transmission.^7-11^ Therefore, both types of asymptomatics contribute significantly to disease transmission. To better control the pandemic of COVID-19, actively discovering, as well as early identifying and quarantining asymptomatics are urgently needed.

Until now, detection of asymptomatic infections has been relied on extensive NAT screening of quarantined individuals. The test sensitivity of NAT depends on the course and the type of clinical COVID-19 syndromes, the collection site, transportation and storage of specimen. About 30% false-negative rates of NAT have been reported in COVID-19 patients.^12^ In particular, recent seroprevalence investigations strongly suggested that COVID-19 cases, especially asymptomatics are greatly underestimated in different countries and regions. SARS-CoV-2 specific IgG response in blood donors reached 3.08% during lockdown of Wuhan city,^13^ consistent with the report of the seropositivity in healthcare workers in Wuhan ranging from 3.2% to 3.8%.^14^ All of these studies indicate that the number of actual infections is at least five times higher than that of the reported cases in Wuhan. In Spain, there was 5% serological positive of national population and 1/3 of these did not report symptoms.^15^ Similar situation occurred in different regions of the United States.^16^ Therefore, these missing asymptomatic cases that are infectious in the community have been substantially overlooked because of the imperfect sensitivity of NAT and passive approaches to discovery them.

The sensitivity of SARS-CoV-2 IgG response can be near 100% when serum samples from COVID-19 patients are acquired within 19 days after first symptoms.^17^ However, asymptomatic individuals had a much longer median duration of viral shedding (19d) than the symptomatic group.^18^ Up to present, humoral immunity in asymptomatic infections with SARS-CoV-2 has not been established. Further assessing dynamics of IgG and IgM responses and neutralizing antibodies to SARS-CoV-2 in asymptomatic COVID-19 infections will help us developing serological tests and improving early identification, understanding the humoral immunity to COVID-19, and providing more rational control strategies for the pandemic.

## Materials

### Definitions

In order to identify and report SARS-CoV-2 infected cases in time, the National Health Commission of China updated the COVID-19 Prevention and Control Plan (3th edition) on Jan 28, 2020, which first emphasized the identification and quarantine of asymptomatic infections. Close contacts with confirmed cases and persons with close social distance during extensive investigation of clusters and tracing infectious sources were required to screen by RT–qPCR. On April 8, 2020, the lockdown had been lifted in Wuhan. Personnel returning to work were also required to screen by RT-qPCR. All NAT positive individuals were asked to provide detail information, including demography, preexisting conditions, exposure history, symptoms, as well as screening records, and accepted centralized isolation for the preceding 14 d. If clinical symptoms and/or lung damage occurred, they should be transferred to the general hospital such as Tongji Hospital for further treatment. NAT negative healthy contacts also complied with home quarantine for 14d. An asymptomatic case was defined as an individual with a positive NAT result but without any relevant clinical symptoms and radiological changes of the lung during quarantine. A mild COVID-19 patient was defined as an individual with nasopharyngeal swabs that were NAT positive for SARS-CoV-2, with symptoms (such as fever, cough or sore throat, etc.) yet without radiological changes of the lung. A close social distance was defined as (1) anyone who had been within approximately 6 feet (2 meters) of a person infected with SARS-CoV-2 for longer than 10 □ min and (2) those who had direct contact with the infectious secretions of a confirmed COVID-19 patient. The National Health Commission of China updated the latest COVID-19 Prevention and Control Plan (7th edition), but continued to use the previous definitions.

### Study design and participants

Between 17 February 2020 and 28 April 2020, 1056 confirmed COVID-19 patients with different severities of illness were continuously recruited from Tongji Hospital, Wuhan, China. Only 51 mild COVID-19 patients who had serial sera with a total of 87 samples and without any preexisting conditions were selected. After screening 11,766 individuals, 63 individuals with asymptomatic infections and 63 healthy contacts without progression were informed in the study. 48 healthy contact and 36 asymptomatic individuals had clear exposure time, respectively. Participants were traced consecutively for 65 days. Serum specimens were collected from each individuals and were stored at −80 °C until use.

### Real-time RT-qPCR

Nasopharyngeal swabs of all participants on enrollment were collected and maintained in viral transport medium. Before detection, all specimens were thermal inactivated in 56°C for 30 min before detection. SARS-CoV-2 infection was confirmed using TaqMan One-Step RT-qPCR Kits (DAAN Gene, Guangzhou, China) which detected ORF1ab and N genes and approved by the China Food and Drug Administration. The RT-qPCR assay kits were performed according to manufacturers’ instructions and the cutoff Ct value was 40 for both genes. The Ct values of both genes were less than 40 was defined as positive.

### SARS-CoV-2 antibody detection

The IgM and IgG antibodies against recombinant nucleoprotein and spike protein of SARS-CoV-2 in serum specimens were detected by commercial kits according to the manufacturer’s instructions (YHLO Biotech, Shenzhen, China). The antibody levels ≥ 10 AU/ mL are reactive (positive), and the results < 10 AU/mL are negative.

### Protein microarray fabrication

The proteins, along with the negative (BSA) and positive controls (anti-Human IgG and IgM antibody), were printed in quadruplicate on PATH substrate slide (Grace Bio-Labs, Oregon, USA) to generate identical arrays in a 2× 7 subarray format using Super Marathon printer (Arrayjet, UK). The microarray was used for serum profiling as described previously with minor modifications. ^19^ Protein microarrays were stored at −80°C until use.

### Microarray-based serum analysis

Proteomic microarray was conducted to probe all the samples.^19^ Briefly, a 14-chamber rubber gasket was mounted onto each slide to create individual chambers for the 14 identical subarrays. The arrays stored at −80°C were warmed to room temperature and then incubated in blocking buffer (3% BSA in 1×PBS buffer with 0.1% Tween 20) for 3 h. Serum samples were diluted 1:200 in PBS containing 0.1% Tween 20, added with 0.1 mg/mL eGFP purified in the same manner as the eGFP tagged proteins and 0.5 mg/mL total *E*.*coli* lysate. A total of 200 μL of diluted serum or buffer only was incubated with each subarray for 2h at room temperature. The arrays were washed with 1×PBST and bound antibodies were detected by incubating with Cy3-conjugated goat anti-human IgG and Alexa Fluor 647-conjugated donkey anti-human IgM (Jackson ImmunoResearch, PA, USA), which were diluted 1: 1,000 in 1×PBST, and incubated at room temperature for 1 h. The microarrays were then washed with 1×PBST and dried by centrifugation at room temperature and scanned by LuxScan 10K-A (CapitalBio Corporation, Beijing, China) with the parameters set as 95% laser power/ PMT 550 and 95% laser power/ PMT 480 for IgM and IgG, respectively. The fluorescent intensity data was extracted by GenePix Pro 6.0 software (Molecular Devices, CA, USA). Signal Intensity was defined as the median of the foreground subtracted by the median of background for each spot and then averaged the triplicate quadruplicate spots for each protein. IgG and IgM data were further analyzed separately. IgG and IgM profiles were built and clustering analysis were conducted to generate heatmaps for overall visualization.

### Neutralization detection using pseudovirus neutralizaion assay

A full length codon-optimized spike protein (S) of the SARS-CoV-2 was first cloned into the lentivirus vector GV367, and then used to generate a eGFP-coexpressing pseudovirus by cotransfected into HEK293T cells with the other two viral packaging help vectors pHelper1.0 and pHelper2.0 (Genechem, Shanghai). 48h after transfection, the supernatants were collected after centrifugation with 4000 g for 10 min at 4□, and further filtrated with 0.45 μm filter. The recombinant pseudovirus were further purified by centrifugation with 25000 rpm for 2 h at 4□ and diluted with PBS. The titer of recombinant pseudovirus was quantified by fluorometry and RT-qPCR. The SARS-CoV-2 pseudovirus neutralization assay was carried out on Vero E6 cells in a 96-well plate. 50 μl serial 2-fold diluted sera from 1:10 to 1:2560 of an individual were prepared, and equal volumes of SARS-CoV-2 pseudovirus were added and the plates were pre-incubated at 37□°C for 1□h. 100 μl of 10^4^ Vero E6 cells were added into each well of a 96-well plate 24h before infection. After washed and added 100μl fresh culture medium, cells were incubated with 100□μl of sera-pseudovirus mixture for 48□h. The cells were collected by digested with 200□μl digestion solution and detected the number of eGFP-expressing cells by FACS. The positive rate of eGFP-expressing cells (PRG) was calculated after collected 1000 cells. Experiments were repeated twice. The neutralization rate (%) was calculated as following:

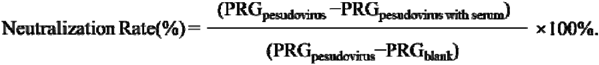

The half-maximal inhibitory concentration (IC50) of each serum sample was determined as the highest dilution ratio of serum with 50% neutralization rate.

### Statistical analysis

All statistical analyses were carried out using SPSS or R software. Signal Intensity of IgM/G response in proteome microarray was defined as the median of the foreground subtracted by the median of background for each spot and then averaged the triplicate spots for each protein. IgM and IgG data were analyzed separately. Log_2_(Singal Intensity) was shown with median (IQR) and Mann–Whitney U test was conducted to test difference between two groups. IC50 was determined using nonlinear regression (SPSS). The Log_2_IC50 was presented with mean (SD) and *t* test was used to test difference between two groups. Cluster analysis was performed with pheatmap package of R. Statistical significance was determined as a value of *p*< 0.05.

## Results

To understand the humoral immunity against SARS-CoV-2 in asymptomatic infections, we first screened by RT-qPCR from 11,776 personnel returning to work, and close contacts with the confirmed cases in different communities of Wuhan by investigation of clusters and tracing infectious sources. Only 12 asymptomatic individuals with positive NAT results were found out. We further conducted a serological survey with the serum samples collected from all participants, using a validated assay for IgM and IgG antibodies against the recombinant antigens containing the nucleoprotein (N) and the spike (S) protein of SARS-CoV-2. Another 51 asymptomatic infections were further discovered because they had positive results for IgG alone, or both IgG and IgM. 63 healthy contacts with both negative results for NAT and antibodies were selected as negative controls. 51 mild patients without any preexisting conditions were also screened from 1056 patients during hospitalization in Tongji Hospital. A total of 177 participants were enrolled in this study and serial serum samples (n=213) were collected (Figure 1). Clear exposure history or days after symptoms onset were obtained from 48 healthy contacts, 36 asymptomatic infections and 51 mild patients. The research was conducted between 17 February 2020 and 28 April 2020. Serum IgM and IgG profiles of 177 participants were further probed using a SARS-CoV-2 proteome microarray. Neutralizing antibody responses in different population were detected by a pseudotyped virus neutralization assay system. The dynamics of IgM and IgG antibodies and neutralizing antibodies were analyzed with exposure time or symptoms onset. Firstly, we found that 63 asymptomatic individuals were classified into four subgroups, based on the results of both NAT and serological tests. 81% (51/63) asymptomatic individuals had negative NAT results but positive IgG responses alone (28/63), or 36.5% (23/63) both IgG and IgM positive. Interestingly, 6.3 % (4/63) were only NAT positive and 12.7% (8/63) were positive for both NAT and IgG (Table 1). Mild patients were further defined as five types. Of mild patients, 25.4% (13/51) were NAT negative but positive IgG alone (4/51), or 17.6% (9/51) both positive for IgG and IgM. 60.3% reported positive results for NAT and IgG alone (10/51), or positive NAT, IgG and IgM (22/51). Importantly, 11.3% (6/51) mild patients were only NAT positive (Table 1).

**Table 1.** Characteristics of study population.

|  | All | Healthy contacts | Asymptomatic(N=63) |  |  |  | Mild(N=51) |  |  |  |  |
| --- | --- | --- | --- | --- | --- | --- | --- | --- | --- | --- | --- |
|  |  |  | Aa(NAT+<br>IgG-IgM-) | Ab(NAT+<br>IgG+IgM-) | Ac(NAT-<br>IgG+IgM-) | Ad(NAT-<br>IgG+IgM+) | Ma(NAT+<br>IgG-IgM-) | Mb(NAT+<br>IgG+IgM-) | Mc(NAT+<br>IgG+IgM+) | Md(NAT-<br>IgG+IgM-) | Me(NAT-<br>IgG+IgM+) |
| N | 177 | 63 | 4 | 8 | 28 | 23 | 6 | 10 | 22 | 4 | 9 |
| Age, years |  |  |  |  |  |  |  |  |  |  |  |
| Mean(SD) | 44.6(19.25) | 43.2(19.85) | 42.8(8.62) | 40.6(10.85) | 46.5(18.05) | 44.7(17.89) | 39.7(25.02) | 39.7(23.77) | 47.6(22.46) | 45.0(31.12) | 54.0(7.40) |
| Median(IQR) | 46(30-59) | 46(27-60) | 41(36-50) | 38(31-52) | 50(29-58) | 40(29-55) | 38(29-46) | 32(20-58) | 51(30-64) | 56(25-66) | 55(48-59) |
| Sex, n(%) |  |  |  |  |  |  |  |  |  |  |  |
| Male | 85(48.0) | 33(52.4) | 0(0.0) | 2(25.0) | 13(46.4) | 10(43.5) | 3(50.0) | 6(60.0) | 11(50.0) | 3(75.0) | 4(44.4) |
| Female | 92(52.0) | 30(47.6) | 4(100.0) | 6(75.0) | 15(53.6) | 13(56.5) | 3(50.0) | 4(40.0) | 11(50.0) | 1(25.0) | 5(55.6) |

IQR, interquartile range; SD, standard deviation.

**Figure. 1.**
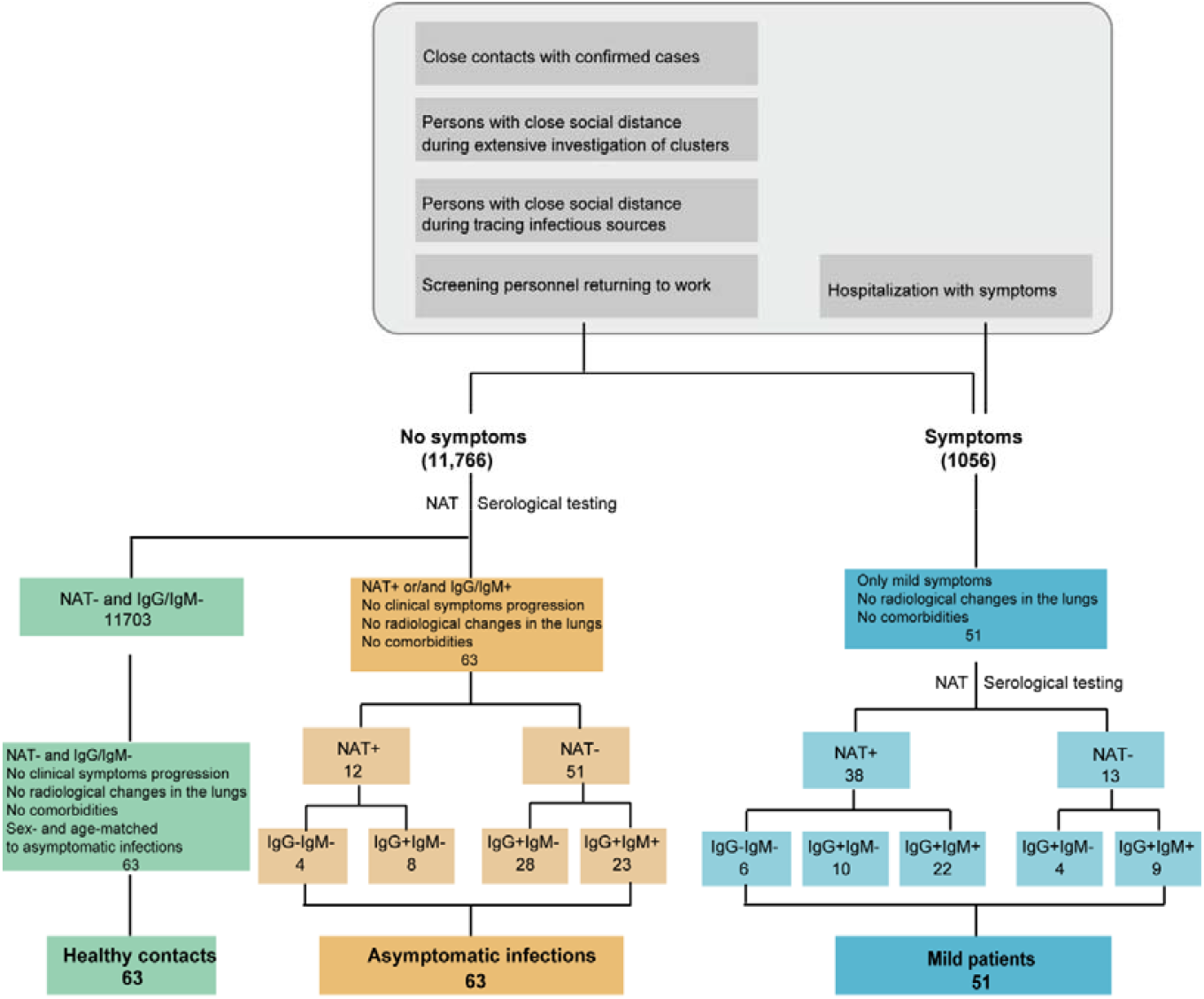
The workflow of screening participants.

To better understand humoral immune response against SARS-CoV-2, IgM (red) or IgG (green) antibody responses to 23 proteins of SARS-CoV-2 were further detected in parallel using a proteome microarray (Figure 2). Overall visualizations of IgM or IgG profiles in 177 participants containing healthy contacts, asymptomatic infections and mild patients were shown in Figure 3 and 4, respectively. Different from the results of clinical diagnosis, healthy contacts, asymptomatic infections and mild cases were not divided into three independent groups, especially based on IgM profiles (Figure 3), which indicated that we cannot distinguish these groups by antibody detection alone. Compared with healthy contacts, both asymptomatic infections and mild cases induced higher IgM (Figure3) and IgG (Figure4) responses, especially against S1, N-protein, N-Nter and N-Cter (Figure 5A and Figure 6A). Asymptomatic infections and mild cases had similar IgM profiles against 23 SARS-Cov2-specific proteins by clustering (Figure 3). However, the levels of IgM responses against S1, N-protein, N-Nter, N-Cter, ORF7b were significant higher in mild patients than asymptomatics (Figure 5A and Figure 6A). Based on IgG profiles, mild patients tended to have stronger IgG response than asymptomatic infections (Figure 4), especially against S1, N-protein, N-Nter, N-Cter and S2-1 (Figure 5B and Figure 6B). And then, we compared the levels of antibodies in each subgroup of asymptomatic infections and mild patients with those of healthy contacts. Except NAT alone positive asymptomatic individuals, other asymptomatic individuals and different subgroups of mild patients induced higher levels of S1, N, N-Nter and N-Cter specific IgM or IgG responses than healthy contacts (Figure 5B and 6B), although these antibody responses could not differentiate the same subgroup between asymptomatic and mild cases (Figure 5C and Figure 6C).

**Figure. 2.**
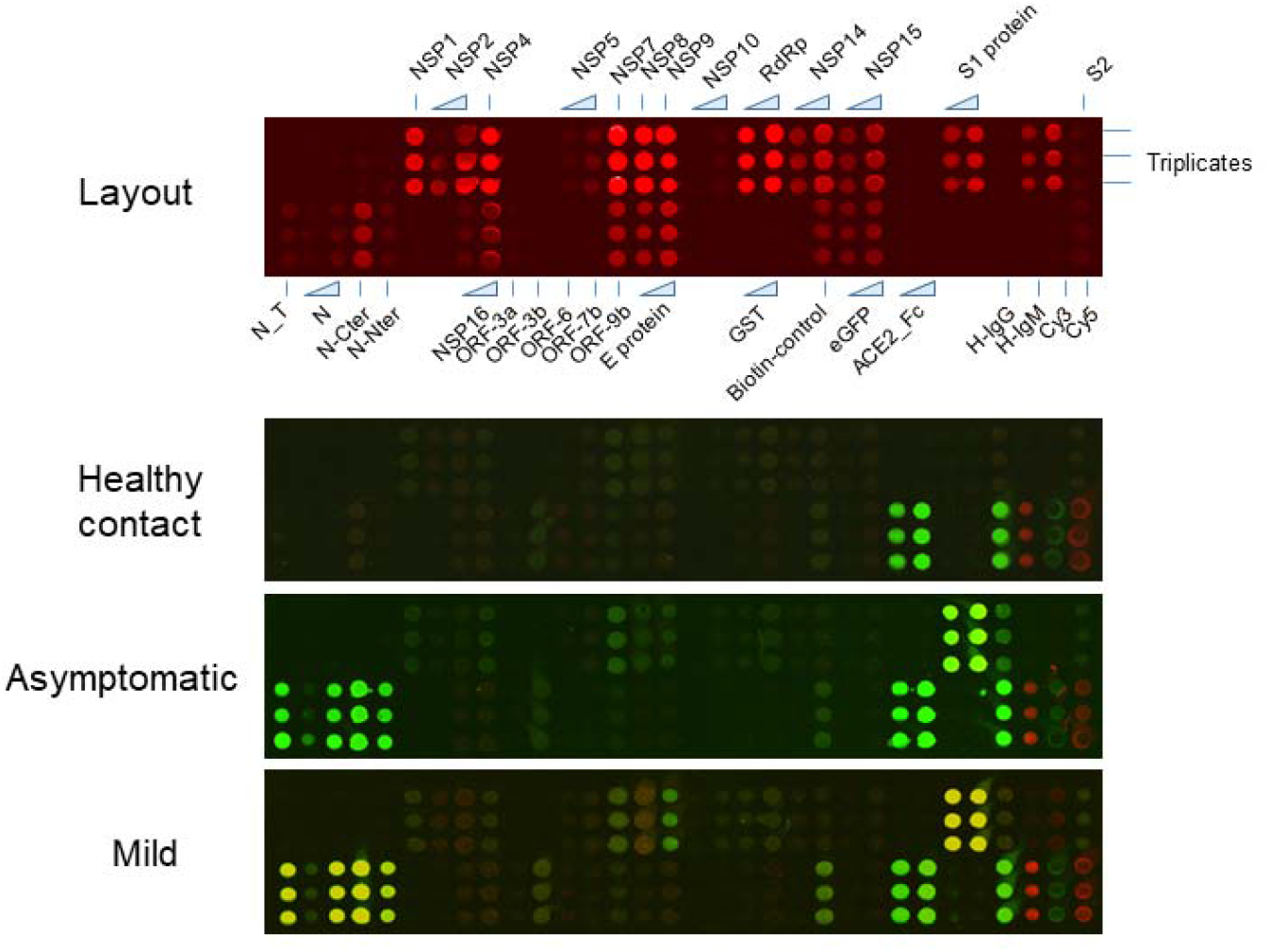
The SARS-CoV-2 proteome microarray layout and serum analysis. (A) There are 23 proteins of SARS-CoV-2 on a single microarray. Each protein is triplicate. The triangles indicate different concentration of one protein. (B) One representative microarray probed with sera of healthy contacts, asymptomatic individuals and mild patients is shown. IgM and IgG responses are shown in red and green, respectively.

**Figure. 3.**
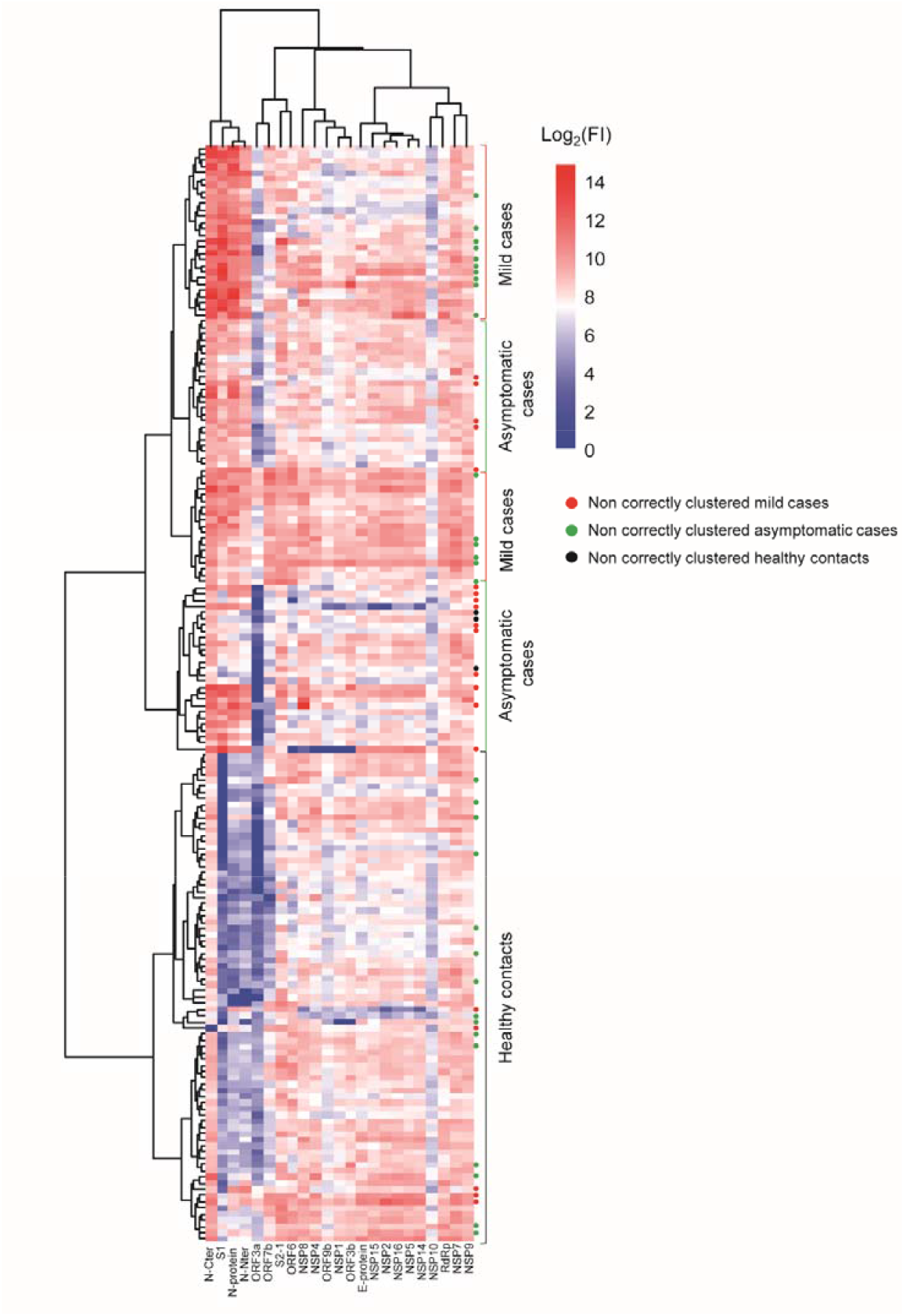
The overall SARS-CoV-2 specific IgM profiles. Heatmap of IgM antibody responses of 63 healthy contacts, 63 asymptomatic individuals and 51 mild patients. Each square indicates the IgM antibody response against the protein (column) in the serum (row). The proteins and sample were arranged with clustering. The red, green and black dots represent non correctly clustered cases in mild, asymptomatic and healthy contacts, respectively. FI:Fluorescence Intensity.

**Figure. 4.**
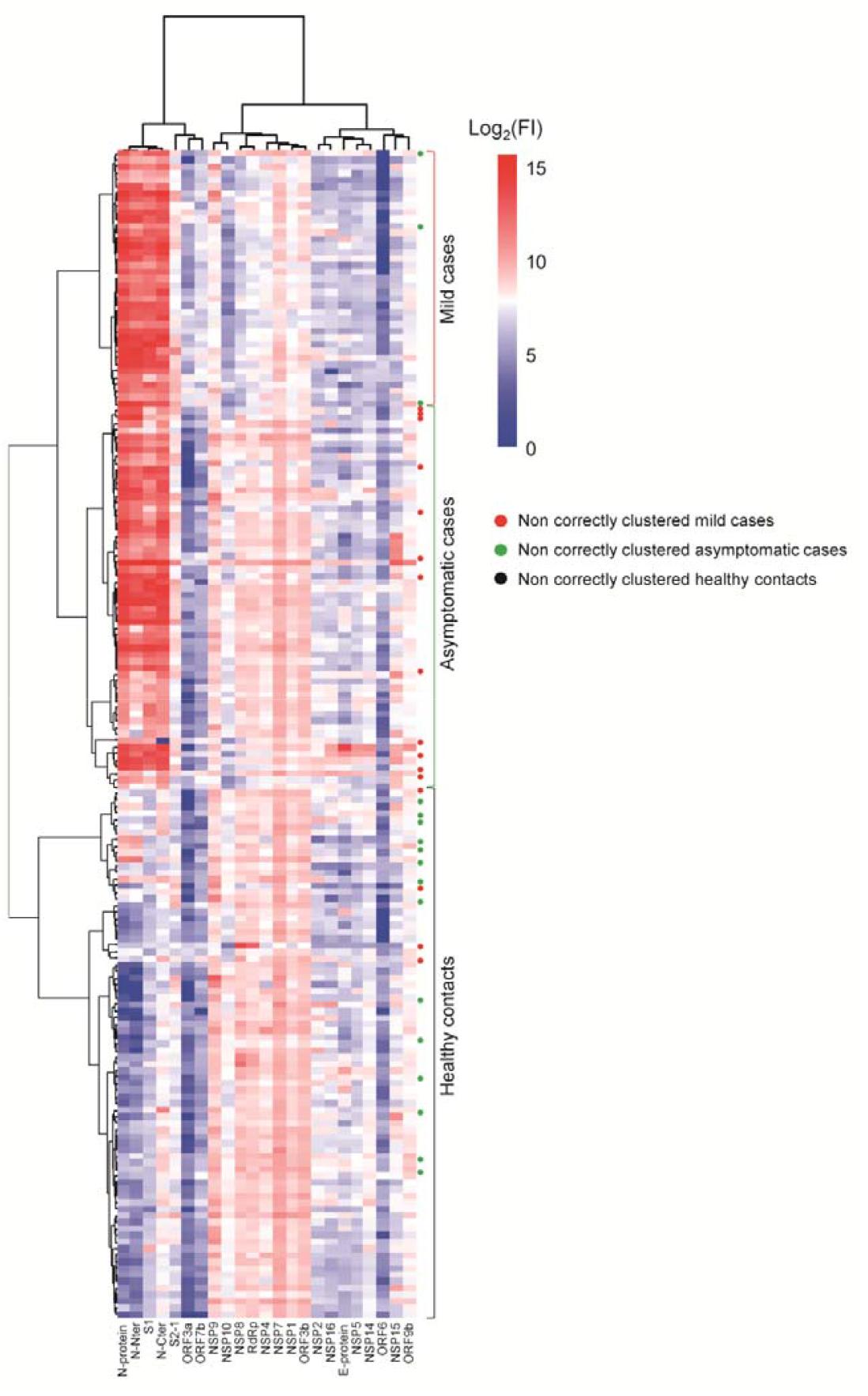
The overall SARS-CoV-2 specific IgG profiles. Heatmap of IgG antibody responses of 63 healthy contacts, 63 asymptomatic individuals and 51 mild patients. Each square indicates the IgG antibody response against the protein (column) in the serum (row). The proteins and sample were arranged with clustering. The red, green and black dots represent non correctly clustered cases in mild, asymptomatic and healthy contacts, respectively. FI: Fluorescence Intensity.

**Figure. 5.**
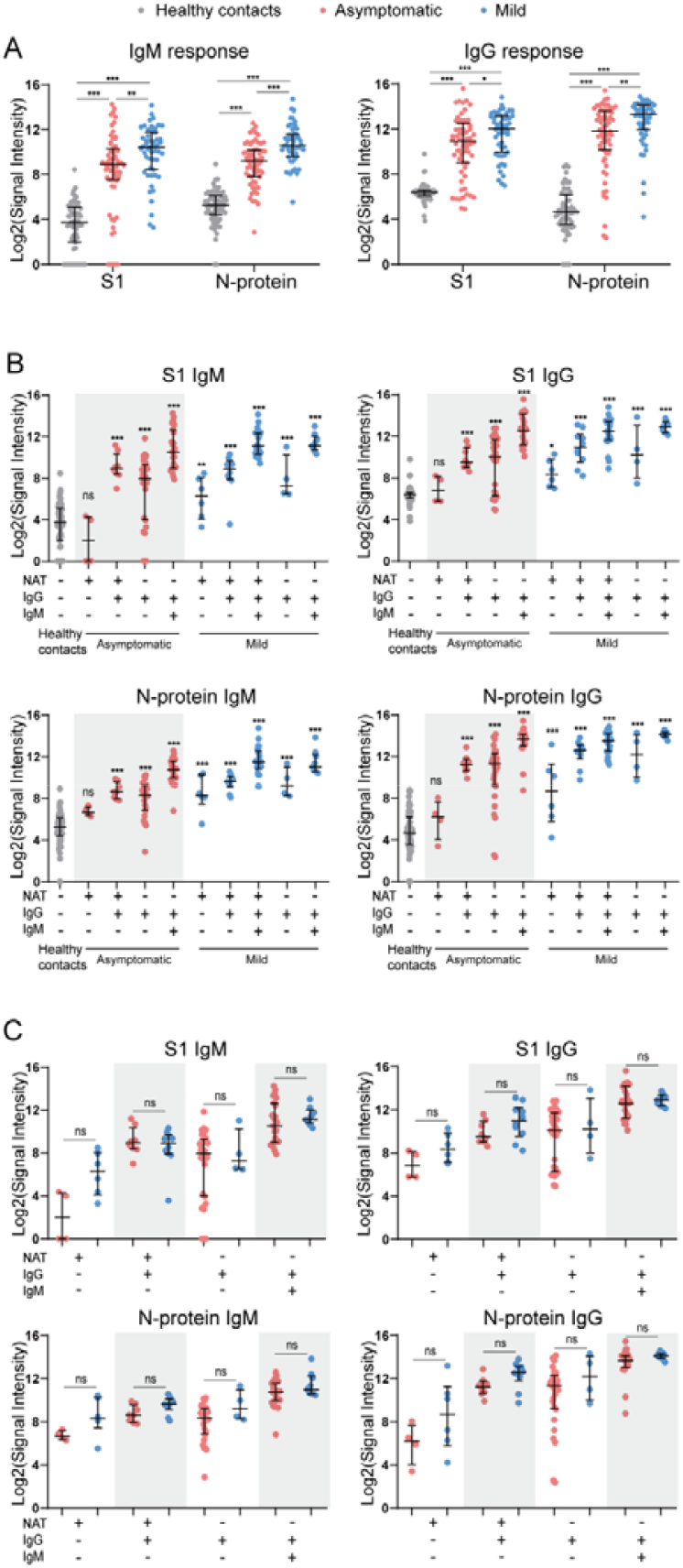
IgM and IgG response to S1 and N proteins in different groups. (A) IgM and IgG response to S1 and N proteins in healthy contacts, asymptomatic individuals and mild patients. (B) Comparison of IgM and IgG responses to S1 and N proteins in different subgroups of asymptomatic individuals and mild patients with that of healthy contacts. (C) Comparison of IgM and IgG response to S1 and N proteins in the same subgroup of asymptomatic individuals and mild patients. Medians and interquartile range value for each group are indicated. Differences between groups were analysed using the Mann–Whitney U test. *** indicated *p* <0.001, ** indicated *p* <0.01, * *p* indicated <0.05, and n.s. indicated not significant.

**Figure. 6.**
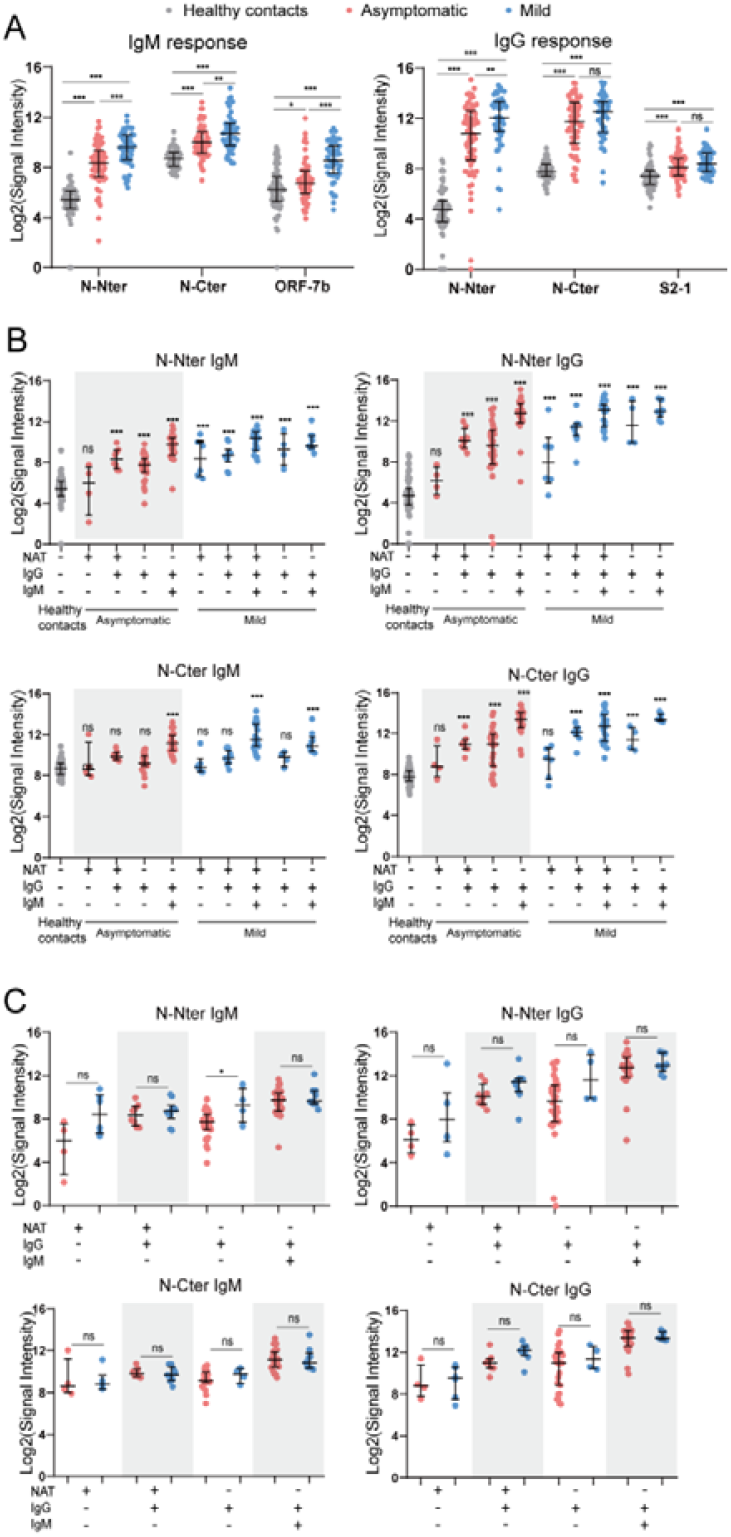
IgM and IgG response to N-Nter, N-Cter and other proteins. (A) IgM response to N-Nter, N-Cter and ORF-7b in healthy contacts, asymptomatic individuals and mild patients (left), IgG response to N-Nter, N-Cter and S2-1 in healthy contacts, asymptomatic individuals and mild patients (right). (B) Comparison of IgM (left) and IgG (right) responses to N-Nter and N-Cter in different subgroups of asymptomatic individuals and mild patients with that of healthy contacts. (C) Comparison of IgM and IgG response to N-Nter and N-Cter in the same subgroup of asymptomatic individuals and mild patients. Medians and interquartile range value for each group are indicated. Differences between two groups were analysed using the Mann–Whitney U test. *** indicated *p* <0.001, ** indicated *p* <0.01, * *p* indicated <0.05, and n.s. indicated not significant.

We further compared the dynamic changes of S1, N, N−Nter, and N-Cter specific IgM and IgG antibodies in 48 healthy contacts and 36 asymptomatic individuals with different exposure time, and in 51 mild patients with days after symptoms onset (Figure 7 and Figure 8). Early to the seventh day after exposure, S1 and N specific IgM and IgG responses were induced in asymptomatic individuals and reached a peak on days from 17d to 25d. Then all of these antibodies began to decline. Except N specific IgG response, other antibodies could not be detectable 2 months after exposure. Interestingly, mild patients had a very distinct dynamic change of antibodies. Early to 1d, S1 specific IgM responses were induced in mild patients and persistently increased until 29 after symptoms onset. Moreover, mild patients elicited higher levels of N specific IgM and IgG responses, which maintained for at least 65 days.

**Figure. 7.**
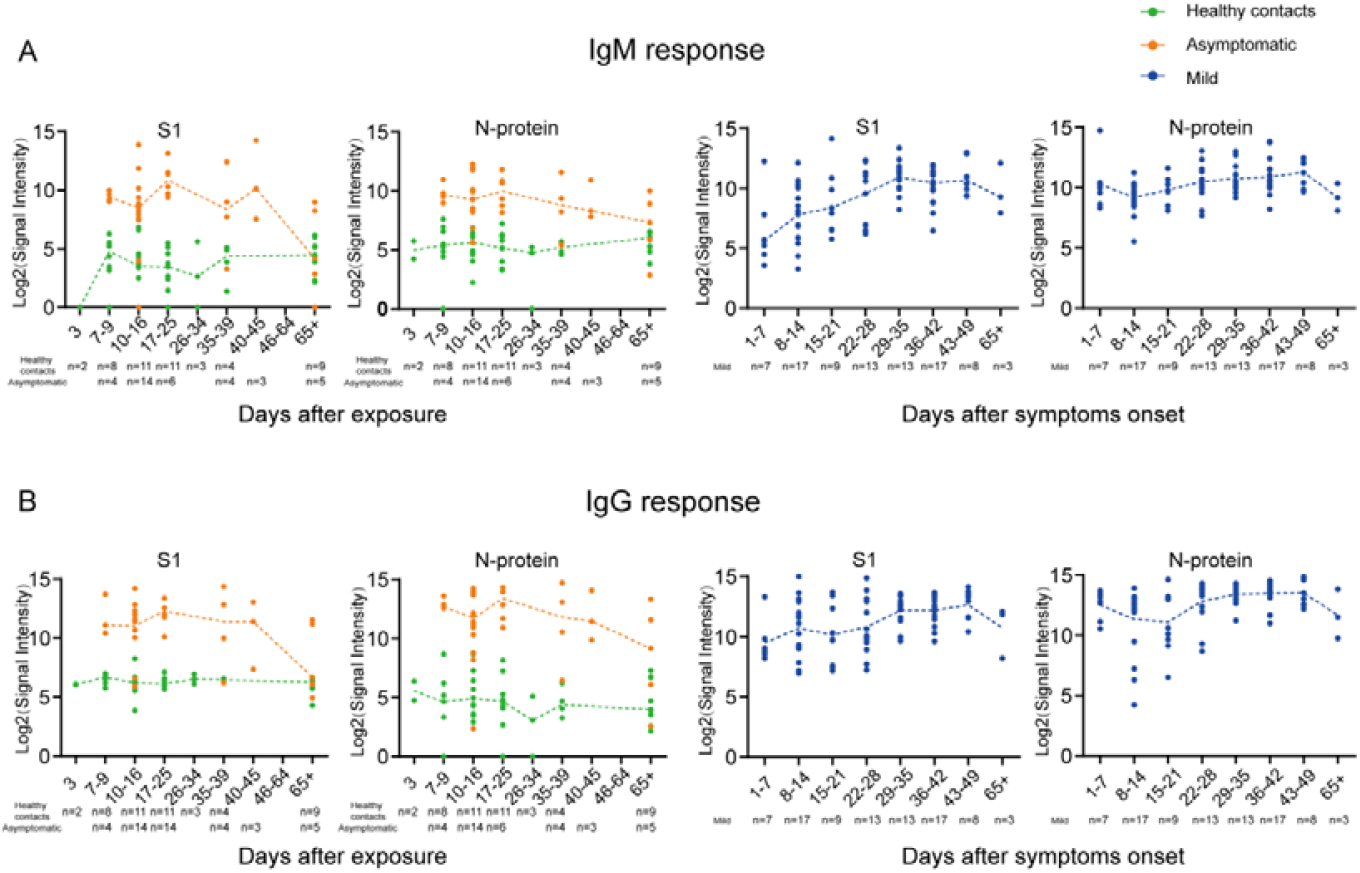
The dynamic change of S1 and N specific IgM and IgG antibody responses. (A) The dynamic change of S1 and N specific IgM antibody responses. (B) The dynamic change of S1 and N specific IgG antibody responses. 48 healthy contacts and 36 asymptomatic individuals with clear exposure time were plotted in sections according to the exposure time. 51 mild COVID-19 patients with serial sera samples (n=87) were segmented according to the symptom onset. The yellow and green line show the levels of antibody responses against SARS-CoV-2 in healthy contacts and asymptomatic at different times after exposure, respectively. The blue line show levels of antibodies against SARS-CoV-2 in mild patients at different times after symptom onset. The bars show medians and interquartile range (IQR).

**Figure. 8.**
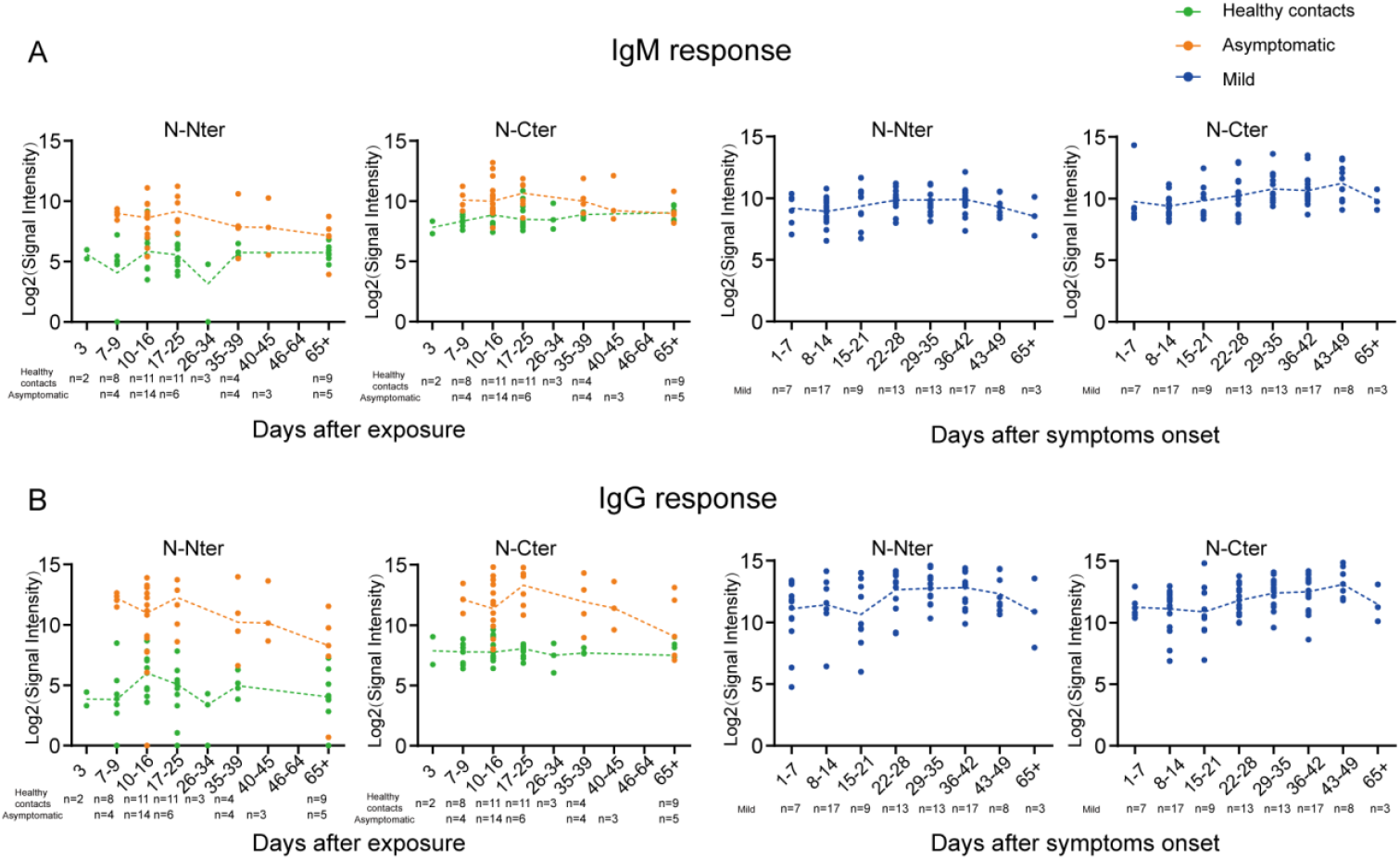
The dynamic change of Nter, N-Cter specific IgM and IgG antibody responses. (A) The dynamic change of Nter, N-Cter specific IgM Antibody responses. (B) The dynamic change of Nter, N-Cter specific IgG Antibody responses. 48 healthy contacts and 36 asymptomatic individuals with clear exposure time were plotted in sections according to the exposure time. 51 mild COVID-19 patients with serial sera partially (n=87) were segmented according to the symptom onset. The yellow and green line in the left show the levels of antibody responses against SARS-CoV-2 in healthy contacts and asymptomatic at different times after exposure, respectively. The blue line in the right show levels of antibodies against SARS-CoV-2 in mild patients at different times after symptom onset. The bars show medians and interquartile range (IQR).

To better understand the role of humoral immunity against infection, we analyzed the dynamic change of the neutralizing antibody responses using a pseudotyped virus based neutralization assay (Figure 9). Interestingly, we found that 36.5% (23/63) asymptomatic individuals, mainly NAT positive (8/12), did not produce neutralizing antibody, 63.5% (40/63) asymptomatic infections and 19% (12/63) healthy contacts only induced low titers of neutralizing antibody, with the mean IC50 1:24 and 1:13, respectively (Figure 8A). Among three groups, mild patients stimulated the highest levels of neutralizing antibody with the mean IC50 1:269. Only 11.8% (6/51) mild patients, mainly NAT alone positive (4/6), did not elicit neutralizing antibody (Figure 9B). More importantly, the neutralizing antibody response in asymptomatic individuals was produced on 7d after exposure and peaked on days from 10d to 25d, then declined with exposure time. However, mild patients produced the neutralizing antibody early to 1d after symptom onset, and the titer rose persistently until 22 days and maintain for at least 65 days (Figure 9C).

**Figure. 9.**
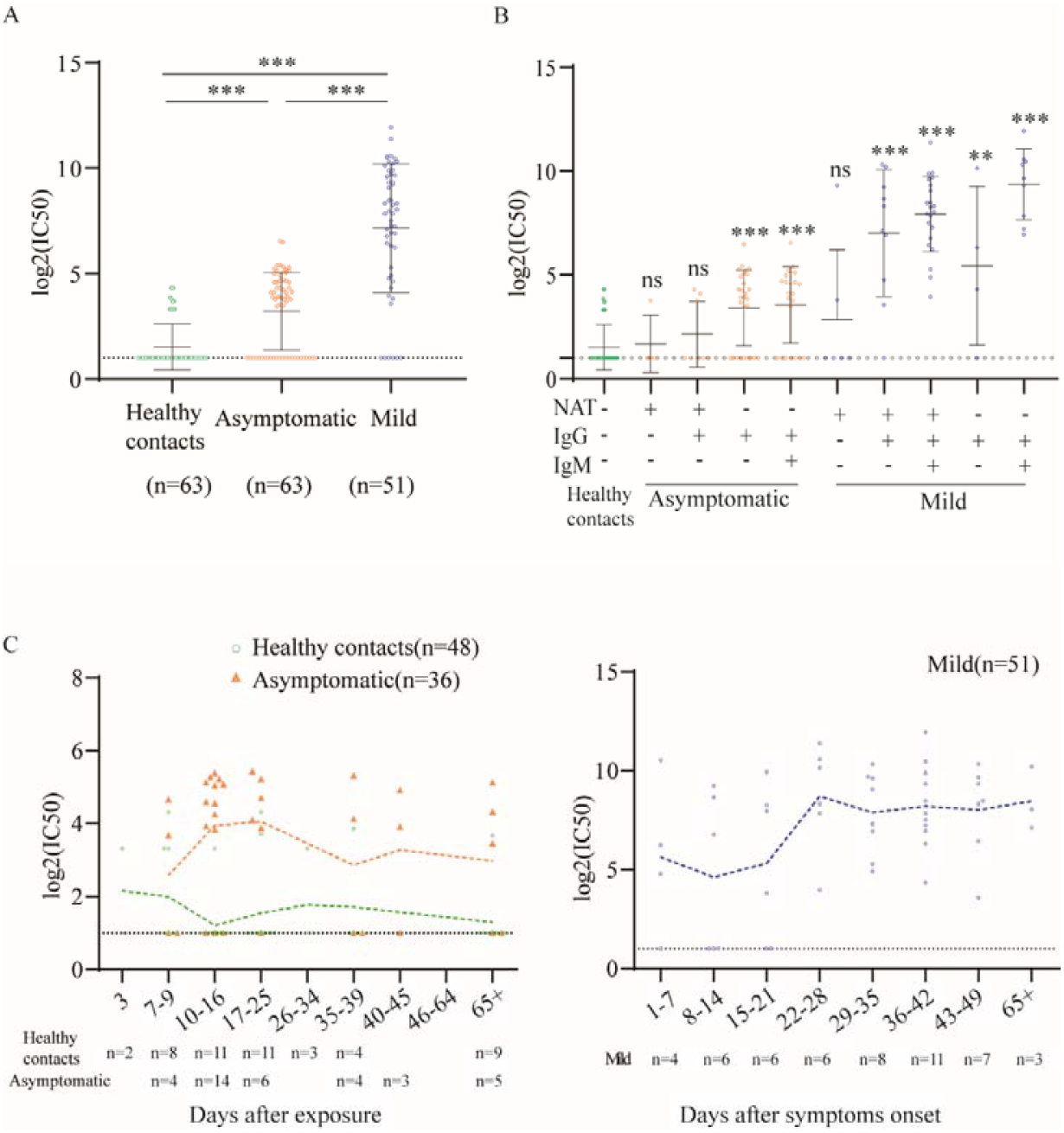
The titers and dynamic change of neutralizing antibody in different groups. (A) Comparison of the titers of neutralizing antibody in different groups. (B) Comparison of the titers of neutralizing antibody in different subgroups of asymptomatic individuals and mild patients with that of healthy contacts. (C) The dynamic change of the titers of neutralizing antibody over time. 48 healthy contacts and 36 asymptomatic individuals with clear exposure time were plotted in sections according to the exposure time. 51 mild COVID-19 patients with serial sera samples were segmented according to the symptom onset. The yellow and green line show the dynamic change of neutralizing ability in healthy contacts(n=48) and asymptomatic individuals (n=36) at different times after exposure, respectively. The blue line shows the dynamic change of neutralizing ability in mild patients at different times after symptom onset. Samples with neutralizing titers below 1:20 were plotted at the dotted line (log_2_IC50=1). Statistical significance in A and B was determined using unpaired t-test and horizontal bars indicate mean and SD. IC50: half-maximal inhibitory concentration.

## Discussion

It remains very difficult to find out asymptomatic COVID-19 infections from healthy population, based on the current control strategy. In this study, five strategies were adopted as recommended by the National Health Commission of China to discover these 63 asymptomatics from more than 10,000 people. Importantly, we firstly demonstrate that only 19% asymptomatic infections had positive NAT results while about 81% detected negative. Our findings are supported by the latest seroprevalence reports that the proportion of asymptomatic infections might be much higher than the reported incidence rate in China,^5,6,13,14^ and also confirm experimentally the view of the prediction model with 87% undiscovered infections.^20^ Combined with the results of seroprevalence investigations in different countries,^15,16^ we can make a conclusion that substantial missing asymptomatic infections cannot be found out in time, based on NAT alone. Therefore, we provide a new perspective that asymptomatic COVID-19 infection should be defined as a person has positive NAT or/and IgM antibody response without clinical symptoms and radiological changes of the lung. This definition will give an additional 36.5% seropositivity of asymptomatics, which is more reasonable and practical than the current used. Under this context, NAT in conjunction with S1 specific IgM serological testing will discover 55.5% asymptomatic infections, except an additional 17.6% positive mild patients. Therefore, this screening strategy will attribute significantly to early identifying and actively discovering infectious source. Asymptomatic individuals had a long duration of viral shedding.^18^ As demonstrated in this study, IgM antibody response against S1 protein can be induced in asymptomatics 1 week after exposure and coincides with the infectious period. Because of rapid emergence and disappearance, S1 specific IgM antibody response might be meaningful to assist NAT for earlier identification of asymptomatic individuals and diagnosis of COVID-19 patients. By contrast, N specific IgG responses persist in the infected host for a longer time and should be more suitable for serological survey and recovery monitor. Therefore, serological testing should be performed based on S or N specific antibody response, not by simultaneously testing responses to S and N antigens of SARS-CoV-2 as performed in current commercial kits. In addition, IgM and IgG profiles against SARS-CoV-2 proteome cannot differentiate the same subgroup of asymptomatic individuals and mild patients, which further indicates the infectiousness of asymptomatic infections. To achieve this end, further investigation to define more accurate serological markers such as profiling B cells epitopes from S1 antigen should be encouraged. Most importantly, the persisting antibodies in asymptomatic individuals only reacted to N, not to S1 antigen. Correspondingly, the titers of neutralizing antibody in asymptomatic individuals were very low and decreased rapidly, consistent the previous report.^18^ These findings indicate that the effectiveness of antibody-mediated immunity could not be used to guarantee the accuracy of an “immunity passport” or “risk-free certificate”. Together these data, our findings might suggest the risks of ‘shield immunity’. Notably, asymptomatic individuals still need immunization with vaccines. Interestingly, strict public health strategies including lockdown, keeping social distance, isolate protection such as wearing mask, washing hands and quarantine were performed in Wuhan, so these undetected asymptomatic individuals did not play important role in disease transmission. However, if these strict public health strategies cannot be implemented, we strongly recommend both NAT and S1 specific IgM antibody response for early identifying, or the latter alone for actively discovering infectious source and that quarantine also should be performed on all IgM positive people. Comparatively, complying with strict public health measures remains the most important strategy to control the pandemic of COVID.

## Data Availability

The data used to support the findings of this study are available from the corresponding author upon request.

## Author Contributors

Dr. Fan had full access to all of the data in the study and takes responsibility for the integrity of the data and the accuracy of the data analysis.

Concept and design: Fan XL, Tao SC

Acquisition, analysis, or interpretation of data: All authors.

Drafting of the manuscript: Fan XL, Lei Q

Critical revision of the manuscript for important intellectual content: All authors.

Statistical analysis: Li Y, Hou HY

Obtained funding: Fan XL

Administrative, technical, or material support: Lai DY, Zhang B

Supervision: Fan XL, Tao SC, Sun ZY

## Ethical approval

The study was approved by the Ethical Committee of Tongji Hospital, Tongji Medical College, Huazhong University of Science and Technology, Wuhan, China (IRB ID:TJ-C20200128). Written informed consent was obtained from all participants enrolled in this study.

